# Early Surgery in Very Preterm Infants is Associated with Brain Abnormalities on Term MRI: A Propensity Score Analysis

**DOI:** 10.1101/2022.09.30.22280582

**Authors:** Katsuaki Kojima, Chunyan Liu, Shelley Ehrlich, Beth M Kline-Fath, Shipra Jain, Nehal A Parikh, the Cincinnati Infant Neurodevelopment Early Prediction Study (CINEPS) Investigators

**Affiliations:** Center for Prevention of Neurodevelopmental Disorders, Perinatal Institute, Cincinnati Children’s Hospital Medical Center, 3333 Burnet Avenue, Cincinnati, OH 45229-3039; Department of Pediatrics, University of Cincinnati College of Medicine, 3230 Eden Avenue, Cincinnati, OH 45267; Division of Biostatistics and Epidemiology, Cincinnati Children’s Hospital Medical Center, 3333 Burnet Avenue, Cincinnati, OH 45229-3039; Department of Radiology, Cincinnati Children’s Hospital Medical Center, 3333 Burnet Avenue, Cincinnati, OH 45229-3039

**Keywords:** Magnetic resonance imaging, neurodevelopmental outcome, newborn, infant, preterm, Bayley-III

## Abstract

**Objective:** To investigate the association between exposure to surgery under general anesthesia during the newborn period and brain abnormalities and neurodevelopmental outcomes in very preterm infants. We hypothesized that exposure to surgery under general anesthesia was associated with structural brain abnormalities on MRI at term-equivalent age and neurodevelopmental impairments at two years corrected age.

**Study design:** This prospective observational study includes 392 infants born at or below 32 weeks’ gestational age. Participants completed brain MRI at term-equivalent age and Bayley-III assessment at two years corrected age. We assessed the MRI global brain abnormality score as our primary outcome. Our secondary outcomes were scores on the Bayley-III Cognitive, Language, and Motor subscales. We evaluated the independent effects of surgery on brain MRI abnormalities and neurodevelopmental outcomes using ANOVA analyses after propensity score matching.

**Results:** All infants completed brain MRI, and 338 (86%) completed neurodevelopmental testing. Forty-five received surgery under general anesthesia. Exposure to surgery was associated with worse MRI abnormalities. None of the developmental outcomes was associated with surgery after propensity score matching. The global brain abnormality score was associated with the Bayley Cognitive and Motor composite scores.

**Conclusions:** Very preterm infants exposed to surgery under general anesthesia were at higher risk of brain abnormalities on MRI at term but not adverse neurodevelopmental outcomes at two years. Global brain abnormality score on MRI was associated with neurodevelopmental outcomes at two years. Brain MRI measures at term may function as sensitive intermediate measures of neurodevelopmental outcomes for very preterm infants.

## INTRODUCTION

Despite advancements in medical care, preterm-born children continue to be at high risk of neurodevelopmental impairments (NDI)^1,2^. Predicting neurodevelopmental outcomes is essential for guiding medical management, counseling parents, and identifying high-risk infants to start early intervention during the critical time window^3^. However, prognosticating preterm infants remains challenging^4^, partly because of the heterogeneous course of events in the NICU, including exposure to surgery under general anesthesia.

Preterm infants receive surgery under general anesthesia for various reasons, including but not limited to necrotizing enterocolitis (NEC), patent ductus arteriosus (PDA), and retinopathy of prematurity (ROP). Surgeries affect the susceptible brains of preterm infants^5^ by inducing systemic inflammation^6^ and through exposure to anesthetic agents^7,8^ and pain^9^ during the procedure. It is challenging to isolate the effects of surgery because infants who receive surgeries are at risk of neurodevelopmental impairments due to the conditions that prompted surgeries, such as NEC^10^. Moreover, socioeconomic status plays a role in neurodevelopment^11^. Appropriate statistical analysis to control for confounding is crucial for understanding the effects of surgery on neurodevelopmental outcomes independent of underlying medical conditions and clinical and social risk factors.

Term brain MRI is a promising tool for predicting or serving as a surrogate measure for neurodevelopmental outcomes in premature infants^12^. However, prior research findings on the effects of surgery on term brain MRI are mixed^13,14^. Furthermore, the relationship between term brain MRI and neurodevelopmental outcomes has not been clearly shown in preterm infants exposed to surgery.

Our aims for this study were twofold. First, we evaluated the potential effects of surgery on neurodevelopment and brain structures in very preterm infants. We hypothesized that exposure to surgery under general anesthesia was associated with neurodevelopmental impairments and structural brain MRI abnormalities independent of clinical and social risk factors. Second, we evaluated the ability of the brain MRI measures at term to function as intermediate sensitive measures of NDI. We hypothesized that the comprehensive and quantitative brain MRI biomarkers at term were associated with neurodevelopmental outcomes.

## METHODS

This is a secondary study of prospectively collected data from the Cincinnati Infant Neurodevelopment Early Prediction Study of very preterm infants. The details of the cohort study are described elsewhere^15^. Briefly, 392 infants born very preterm (born at or before 32 weeks of gestational age) were recruited from 5 neonatal intensive care units (NICUs) in the greater Cincinnati area. All infants born very preterm in one of these NICUs between September 2016 and November 2019 were eligible for inclusion. Infants were excluded if they met any of the following criteria: (1) known chromosomal or congenital anomalies affecting the central nervous system; (2) cyanotic heart disease; or (3) hospitalization and mechanical ventilation with greater than 50% supplemental oxygen at 45 weeks postmenstrual age (PMA). The Cincinnati Children’s Hospital institutional review board approved the study, and the review boards of the other participating hospitals approved the study based on an established reliance agreement. Written informed consent was provided by a parent or guardian of each study infant after they were given at least 24 hours to review the consent and ask questions to the investigators.

Trained research staff collected a predefined list of maternal characteristics, pregnancy/delivery data, and infant data, including the type, number, and duration of surgical procedures^15^. Infants were categorized into either the surgery group (if they had undergone a surgical procedure under general anesthesia before discharge) or the non-surgery group. For this study, we excluded infants from the analysis if they received surgery after term MRI. Potential confounders of the association between exposure to surgery under general anesthesia and neurodevelopmental outcomes included gestational age at birth, birth weight, sex, antenatal steroid exposure, low 5-minute APGAR score, exposure to expressed breast milk in the first 28 days, maternal infection, chorioamnionitis, limited maternal education (<12^th^ grade), low income (< $40,000), maternal non-private medical insurance, early intraventricular hemorrhage or cerebellar hemorrhage, proven NEC, and the Clinical Risk Index for Babies – second edition (CRIB-II) score^16^. CRIB-II is a clinically validated score associated with neurodevelopmental outcomes^17^ and term brain MRI abnormalities^18,19^. It is calculated using five perinatal risk factors: birth weight, gestational age, sex, admission temperature, and serum base deficit. Baseline maternal and neonatal characteristics significantly associated with both exposures to surgery and outcome variables (p < .05) were included in the propensity score calculation.

MRI of the brain was obtained between 39 and 45 weeks of PMA. Images were collected during natural sleep with a 3T Philips Ingenia scanner (Philips Healthcare) and a 32-channel head coil located at Cincinnati Children’s Hospital, as previously described^20^. We used our feed-and-wrap technique to avoid the use of sedation. We evaluated the total brain tissue volume, four MRI metrics of cortical maturation (surface area, sulcal depth, gyrification index, and curvature), and a single composite MRI abnormality severity score (global brain abnormality score), as described by Kidokoro et al.^21^. We used the automated developing Human Connectome Pipeline (dHCP) to process T2-weighted MRI scans^22^; dHCP uses cortical and subcortical segmentation algorithms to calculate cortical maturation metrics such as sulcal depth, gyrification index, white matter curvature, and cortical surface area. A value for each cortical metric is generated for all regions of the Gousias brain atlas^23^, merged lobar regions, and the full brain. Total brain tissue volume was calculated by subtracting total cerebrospinal fluid from total brain volume. After running the dHCP pipeline algorithm, we visually inspected the results. Participants with moderate or severe ventriculomegaly (VM) or poor-quality imaging (poor tissue identification, improper segmentation, or artifact) were excluded from analyses using the dHCP algorithm. However, these infants were included in analyses for the global brain abnormality score, which accounts for macroscopic intracranial injuries, such as intraventricular hemorrhage, periventricular leukomalacia, and VM, in their evolved form.

The global brain abnormality score is defined by signal abnormalities and impaired brain development for cerebral white matter, cortical gray matter, deep nuclear gray matter, and cerebellar abnormalities^21^. All MRI scans and two-dimensional biometric measurements were obtained by a single pediatric neuroradiologist to yield a global brain abnormality score, as described by Kidokoro et al.^21^ and previously published by our group^24^. Global brain abnormality scores were log-transformed before the analysis to correct its skewed distribution. Back-transformed results were reported.

Skilled examiners performed developmental assessments at 22-26 months corrected age using the Bayley Scales of Infant & Toddler Development 3^rd^ edition (BSID-III)^25^. All Bayley examiners were trained using the NICHD Neonatal Research Network standards. The cognitive subscale of the BSID-III examines global mental function derived from memory, object manipulation, and/or problem-solving; the language subscale assesses language understanding and vocabulary; and the motor subscale examines fine and gross motor function. We examined the composite scores of each of these subscales, standardized to a mean score of 100, a standard deviation of ± 15, and a range of 40 - 160^25^. Bayley testers were blinded to cortical maturation indices and global brain abnormality scores relevant to this study but not clinical structural neuroimaging results.

Patient demographics were summarized using mean and standard deviations or frequency and percentages. MRI outcomes and Bayley-III scores were summarized using median and interquartile ranges. The associations between the potential confounders and the exposure to surgery were examined by univariate analysis using the Wilcoxon rank sum test for continuous variables and Chi-square test or Fisher’s exact test for categorical variables. Antecedent variables significantly associated with exposure to surgery (p-values < 0.05) were then tested for associations with each of the neurodevelopmental outcomes (Bayley Cognitive, Language, and Motor scores) using 1-way ANOVA or linear regression. The union of the variables associated with at least one neurodevelopmental outcome (p-values < 0.05) were used as confounders for the subsequent propensity score model. Baseline characteristics were compared between the participants who completed the study and those lost to follow-up.

Propensity score analysis is a statistical method that balances the baseline characteristics between cases and controls^26^. The propensity score is a summary score measuring how likely each participant will be in the case group, given their baseline characteristics. The propensity score model was fitted using all the variables associated with surgery and at least one of the neurodevelopmental outcomes. The calculated propensity scores were then inspected in three different ways. First, propensity scores were used for 1:1 matching of the surgery group and non-surgery group. Each infant in the surgery group was matched with one in the non-surgery group based on the propensity score. Propensity scores of the paired infants were within 20% SD caliper. Second, we matched infants in the surgery and non-surgery groups using a flexible matching ratio between 1:10 and 10:1^27^. Propensity scores of matched infants were within 20% SD caliper. We call this M: N full matching. Lastly, we calculated inversed probability weight for all infants and included all participants in the analysis. Weight was calculated using the following formula.

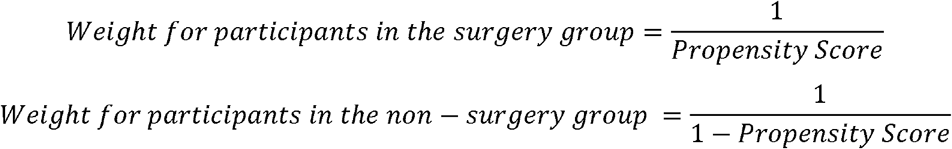

We created density plots comparing the surgery and non-surgery groups to evaluate the overlap of baseline characteristics between the groups after the above three analysis methods. We determined which propensity score analysis method to use based on the overlap between the groups and the number of samples included in the analysis. Association between exposure to surgery and outcome measures (MRI outcomes and Bayley-III scores) was assessed by univariate analysis using the ANOVA analysis after propensity score matching.

Association between MRI outcomes and Bayley-III scores in all the study participants was assessed using multivariable regression analysis. We did not use propensity score matching for this analysis because we included all the participants regardless of exposure to surgery. In the regression model, all the potential confounders and exposure to surgery were included as covariates. We created separate linear regression models to evaluate the relationship between the number of surgeries and neurodevelopmental outcomes.

## RESULTS

Of the original cohort of 392 infants, brain MRI was performed on all 392 infants at term-equivalent age. Neurodevelopmental testing was performed in 338 infants (86.2%) at two years corrected age. Forty-five infants (11.5%) had at least one surgical procedure under general anesthesia before being discharged from the NICU. Most infants (28 infants, 62.2%) received only one surgery. Eight (17.8%), five (11.1%), and four (8.9%) infants received 2, 3, and 4 or more surgeries, respectively. The median cumulative surgery duration was 187 minutes (range: 48 - 838 minutes) in 44 infants with available data. Twenty-five infants (55.6%) received gastrointestinal surgery, such as gastrostomy tube placement and laparotomy; 15 (33.3%) received pulmonary procedures, such as bronchoscopy; nine (20.0%) received cardiac surgery, including PDA closure; and two (4.4%) received laser surgery for ROP. Four infants received surgery after their study brain MRI. Therefore, these four infants were excluded from the analysis using brain MRI findings as an outcome. We compared baseline characteristics of infants evaluated at follow-up (n = 338) with those who were lost to follow-up (n = 54) and found that these two cohorts were similar (*p* > .05) except for gestational age, birth weight, sex, CRIB-II score, and insurance (**Table 1; available at www.jpeds.com**). Those lost to follow-up had favorable clinical profiles (higher gestational age, lower CRIB-II scores) but adverse socioeconomic backgrounds (fewer private insurance).

**Table 1:** Distribution of important clinical and socioeconomic factors between those who were evaluated at follow-up and lost to follow-up

| **Clinical and socioeconomic factors<sup>\*</sup>** | **Evaluated at follow-up (n = 338)** | **Lost to follow-up (n = 54)** | **p value** |
| --- | --- | --- | --- |
| Gestational age at birth, weeks, median (range) | 29.0 (23.0-33.0) | 30.5 (23.0-33.0) | .015 |
| Birth weight, grams, mean ± SD | 1269 ± 436 | 1444 ± 501 | .024 |
| Female sex | 164 (49%) | 18 (33%) | .038 |
| Antenatal steroid exposure | 312 (92%) | 49 (91%) | .600 |
| Apgar scores <5 at 5 minutes<sup>†</sup> | 42 (13%) | 6 (11%) | .762 |
| Exposure to maternal milk in first 28 days | 132 (39%) | 22 (41%) | .814 |
| Maternal infection<sup>‡</sup> | 48 (14%) | 8 (15%) | .938 |
| Moderate-severe histological chorioamnionitis or clinical chorioamnionitis<sup>§</sup> | 50 (15%) | 5 (9%) | .277 |
| Intraventricular hemorrhage grade 2, 3, or 4, or cerebellar hemorrhage on early cranial ultrasound | 33 (10%) | 4 (7%) | .582 |
| CRIB II score, mean ± SD | 6.3 ± 4.0 | 4.8 ± 3.7 | .008 |
| Maternal education ≤12<sup>th</sup> grade<sup>¶</sup> | 18 (5%) | 3 (6%) | .747 |
| Annual household income < \$40,000 | 148 (46%) | 26 (54%) | .263 |
| Maternal non-private medical insurance | 168 (50%) | 39 (72%) | .002 |
| Necrotizing enterocolitis | 17 (5%) | 2 (4%) | 1.00 |

\*All values are N (%) unless otherwise noted.
†Data missing for 4 infants (evaluated at follow-up 4).
‡Data missing for 5 infants (evaluated at follow-up 5).
§Data missing for 7 infants (evaluated at follow-up 6, lost to follow-up 1).
¶Data missing for 11 infants (evaluated at follow-up 7, lost to follow-up 4).
||Data missing for 19 infants (evaluated at follow-up 13, lost to follow-up 6).

Infants in the surgery group had lower gestational age and birth weight than the non-surgery group (**Table 2**). They more frequently had low 5-minute Apgar scores, chorioamnionitis, intraventricular hemorrhage on early cranial ultrasound, NEC, higher CRIB-II scores, and non-private maternal insurance than the non-surgery group. All these variables were also significantly associated with at least one of the neurodevelopmental outcomes. We used these variables to calculate propensity scores. Gestational age and birth weight were not included in the propensity score calculation because the CRIB-II score includes these variables.

**Table 2:** Distribution of important clinical and socioeconomic factors at baseline between surgery and non-surgery groups

| **Clinical and socioeconomic factors\*** | **Surgery (n = 45)** | **Non-surgery (n = 347)** | **p value** |
| --- | --- | --- | --- |
| Gestational age at birth, weeks, median (range) | 27.0 (23.0-33.0) | 30.0 (23.0-33.0) | < .0001 |
| Birth weight, grams, mean ± SD | 1089 ± 513 | 1319 ± 434 | .0002 |
| Female sex | 20 (44%) | 162 (47%) | .777 |
| Any antenatal steroid exposure | 40 (89%) | 321 (93%) | .380 |
| Apgar scores <5 at 5 minutes<sup>†</sup> | 13 (30%) | 35 (10%) | .0002 |
| Exposure to maternal milk in first 28 days | 15 (33%) | 139 (40%) | .385 |
| Maternal infection<sup>‡</sup> | 6 (14%) | 50 (15%) | .919 |
| Moderate-severe histological chorioamnionitis or clinical chorioamnionitis<sup>§</sup> | 14 (33%) | 41 (12%) | .0003 |
| Intraventricular hemorrhage grade 2, 3, or 4, or cerebellar hemorrhage on early cranial ultrasound | 16 (36%) | 21 (6%) | <.0001 |
| CRIB II score, mean ± SD | 8.9 ± 4.5 | 5.7 ± 3.8 | <.0001 |
| Maternal education <12<sup>th</sup> grade<sup>¶</sup> | 0 (0%) | 21 (6%) | .149 |
| Annual household income < \$40,000 | 21 (50%) | 153 (46%) | .644 |
| Maternal non-private medical insurance | 30 (67%) | 177 (51%) | .048 |
| Necrotizing enterocolitis | 13 (29%) | 6 (2%) | <.0001 |

\*All values are N (%) unless otherwise noted.
<sup>†</sup>Data missing for 4 infants (surgery 1, non-surgery 3).
<sup>‡</sup>Data missing for 5 infants (surgery 2, non-surgery 3).
<sup>§</sup>Data missing for 7 infants (surgery 2, non-surgery 5).
<sup>¶</sup>Data missing for 11 infants (surgery 2, non-surgery 9).
<sup>||</sup>Data missing for 19 infants (surgery 3, non-surgery 16).

The distribution of the calculated propensity scores was compared in three ways. First, 1:1 matching within 20% SD caliper resulted in 36 pairs. Second, we performed M: N full matching within a 20% SD caliper using the case to control matching ratios ranging from 10:1 to 1:10. This resulted in 185 matched sets. Finally, we used inversed probability weighting (IPW). The density plots of the propensity scores after these three methods (**Figure 1; available at www.jpeds.com**) showed the best overlap between surgery and non-surgery groups in the 1:1 matched data, followed by full-matched data and IPW. However, the number of samples included in the analysis was in the reverse order, with the smallest sample size in 1:1 matching (n = 72; 36 pairs), followed by full-matching (n = 179) and IPW (n = 383). Thus, we used full matching data for our outcome analysis to balance sample size and matching quality.

**Figure 1.**
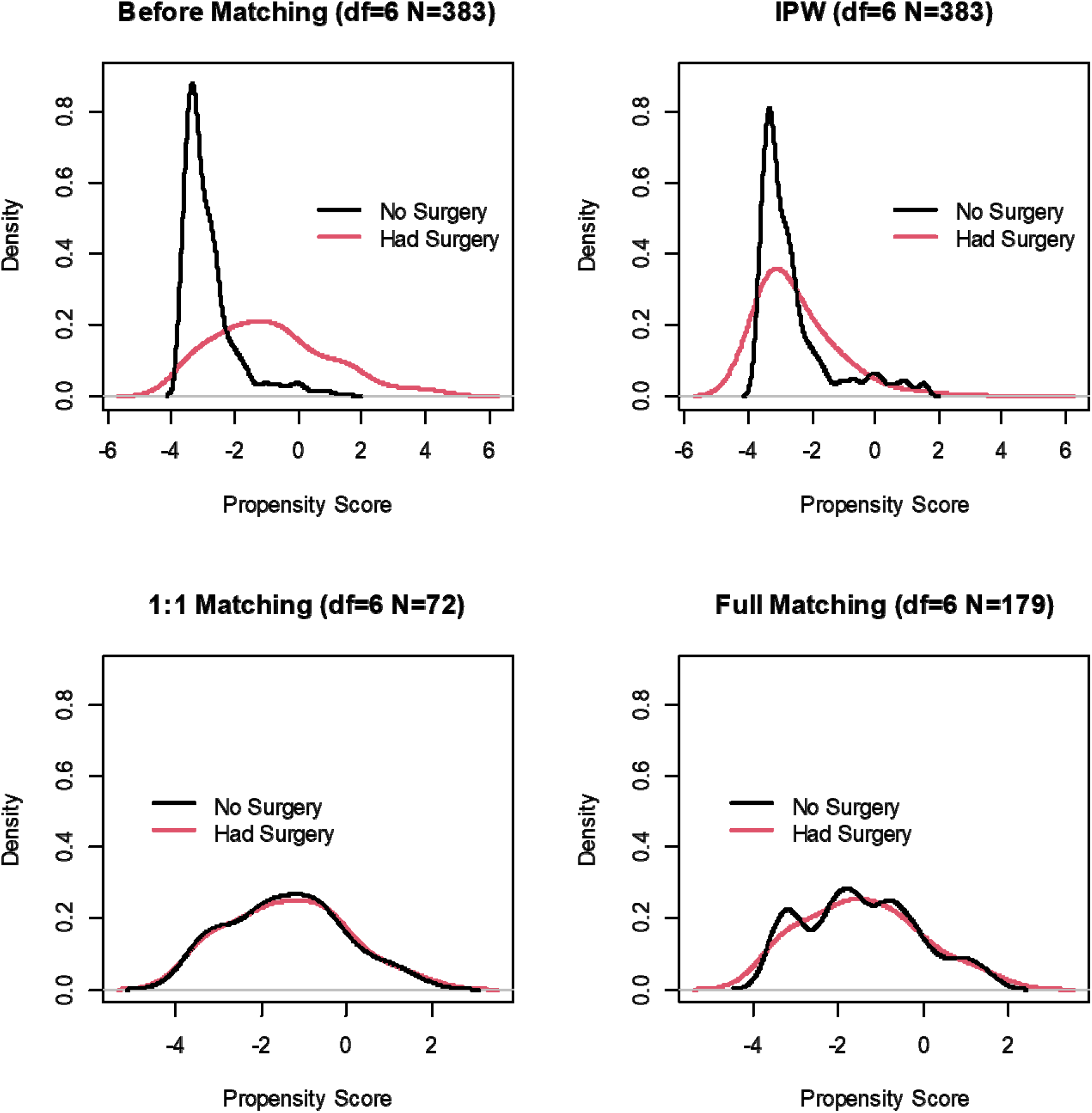
Density plot of propensity scores by surgery groups. Top-Left panel: before matching; Top-Right panel: Inversed probability weighting (IPW) method; Bottom-Left panel: after 1:1 matching using caliper of 20% SD (N = 72, 36 pairs); Bottom-Right panel: after M: N full matching (N=179).

The ANOVA analyses using full-matched data showed that the exposure to surgery was associated with worse MRI measures at term, including higher global brain abnormality score (*p* < .0001), smaller total brain tissue volume (*p* < .0001), and lower surface area (*p* = .001) (**Table 3**). Univariate analyses showed that exposure to surgery was significantly associated with lower composite scores of all the neurodevelopmental outcome measures (**Table 4; available at www.jpeds.com**). However, no two-year developmental outcomes were associated with surgery exposure after propensity score matching (**Table 3**). The Bayley-III Motor composite score showed a trend towards significance (*p* = .100). None of the Bayley composite scores was associated with the number of surgeries (*p* > .21).

**Table 3:**
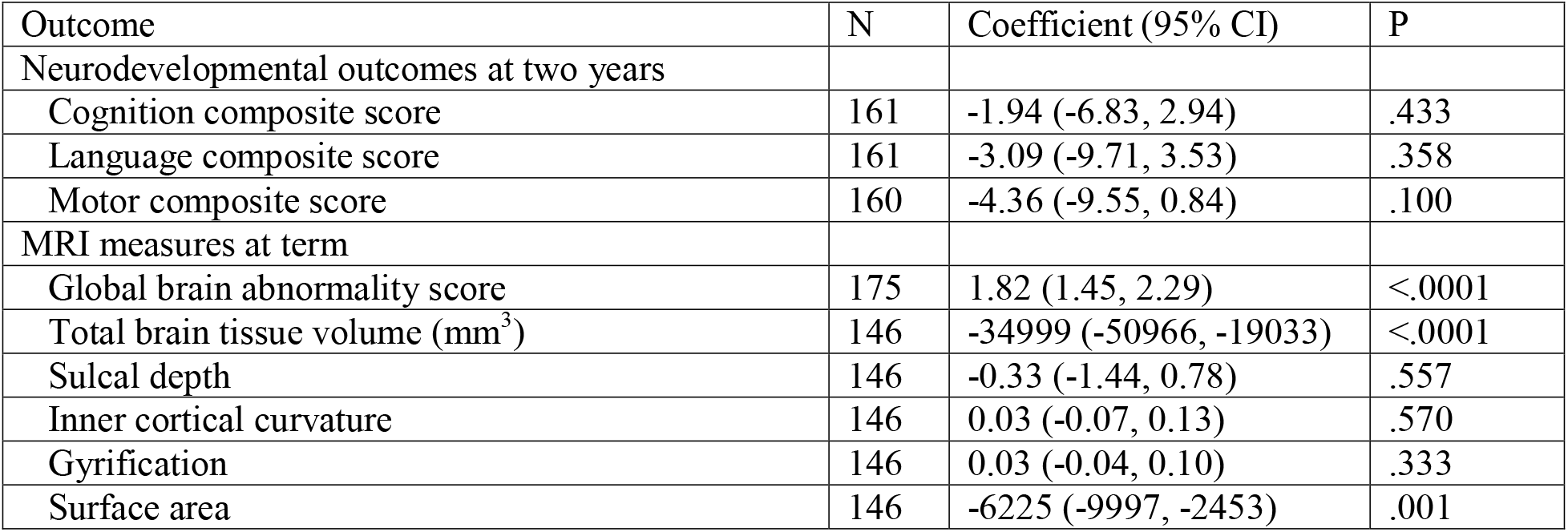
ANOVA analyses examining the association between exposure to surgery and several term MRI and neurodevelopmental outcome measures after propensity score matching

**Table 4:** Univariate analyses showing the effects of exposure to surgery on outcome measures

| Outcome* | Surgery | Non-surgery | P† |
| --- | --- | --- | --- |
|  | N = 41 | N = 347 |  |
| Global brain abnormality score | 12 (8, 14) | 4 (2, 7) | <.0001 |
|  | N = 27 | N = 308 |  |
| Total brain tissue volume | 357306 (329007, 377094) | 398741 (366039, 428255) | <.0001 |
|  | N = 27 | N = 307 |  |
| Sulcal depth | 24.2 (21.8, 26.1) | 25.9 (23.5, 27.9) | .005 |
|  | N = 27 | N = 307 |  |
| Inner cortical curvature | 2.1 (2.0, 2.3) | 2.1 (2.0, 2.2) | .600 |
|  | N = 27 | N = 307 |  |
| Gyrification index | 2.5 (2.4, 2.7) | 2.5 (2.4, 2.6) | .818 |
|  | N = 27 | N = 307 |  |
| Surface area | 79685 (73215, 89114) | 89734 (80804, 96995) | .001 |
|  | N = 40 | N = 298 |  |
| Cognitive composite score | 85.0 (70.0, 95.0) | 95.0 (85.0, 100.0) | .002 |
|  | N = 40 | N = 294 |  |
| Language composite score | 84.5 (59.0, 98.5) | 94.0 (79.0, 106.0) | .004 |
|  | N = 39 | N = 296 |  |
| Motor composite score | 88.0 (73.0, 94.0) | 94.0 (88.0, 100.0) | .0002 |

\*All values are median (interquartile ranges).
†Wilcoxon Rank Sum Test

The global brain abnormality score was significantly associated with Bayley cognitive composite scores (*p* = .006) and motor scores (*p* = .044) but not with language scores (*p* = .108) in multivariable models (**Table 5**). None of the other five secondary MRI measures were associated with neurodevelopmental outcome measures.

**Table 5:**
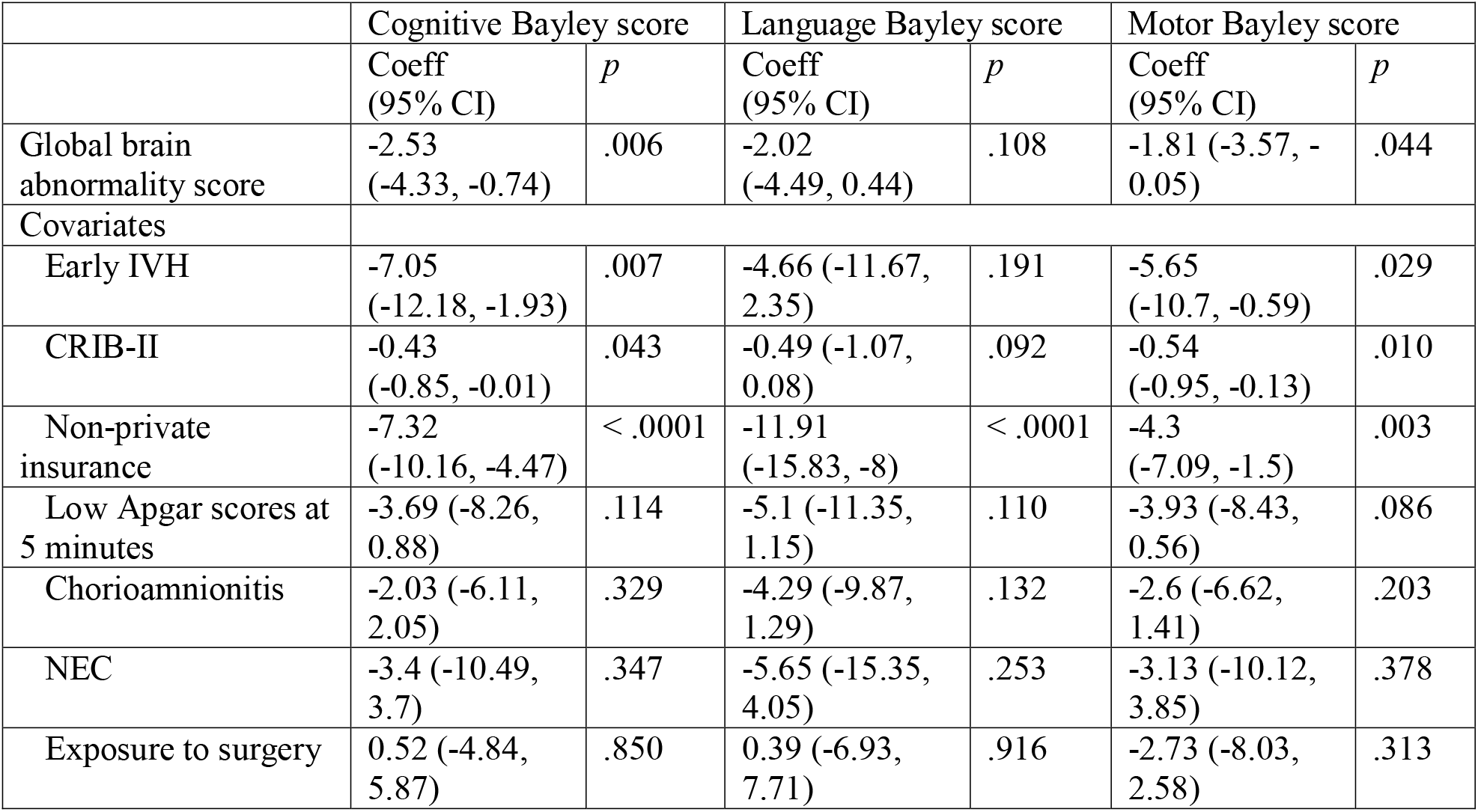
Multivariable models evaluating the association between global brain abnormality score at term and neurodevelopmental scores at two years corrected age.

## DISCUSSION

Studying a well-characterized and geographically defined cohort of infants born very preterm, we showed exposure to surgery under general anesthesia before NICU discharge was associated with worse brain MRI measures at term than for very preterm infants with similar clinical and social risk factors but without surgical exposure. In univariate analysis, exposure to surgery under general anesthesia was associated with worse neurodevelopmental outcomes in very preterm infants. However, this association was no longer present after adjusting for baseline risk factors and other confounders using propensity score matching. Global brain abnormality scores in term brain MRI were associated with neurodevelopmental outcomes at two years old after controlling for clinical and social risk factors, including exposure to surgery.

Our findings are clinically significant because we showed the potential clinical utility of term brain MRI global brain abnormality score as a sensitive intermediate biomarker associated with neurodevelopmental outcomes of very preterm infants. Global brain abnormality score on MRI at term could support clinical management by identifying infants at high risk for neurodevelopmental impairments and benefit from early intervention. We also showed that surgery under general anesthesia poses risks of adverse brain structural development in very preterm infants. This information may help counsel the risks and benefits of surgeries, though some surgeries may be inevitable, and the decision to undergo surgery may not be affected. Our non-significant findings associating surgery with neurodevelopmental scores may have been underpowered and support the use of shorter-term neuroimaging measures as more sensitive intermediate biomarkers of neurodevelopment. The global brain abnormality score could serve as such a surrogate measure for neurodevelopmental outcomes in future trials of surgical interventions in preterm infants.

Our results affirmed a prior study^13^ that showed an association between exposure to surgery and more severe brain abnormalities at term in very preterm infants. Filan et al. studied 227 very preterm infants, of whom 30 received surgery, and showed more frequent white matter injuries and smaller total brain tissue volume in the surgery group^13^. However, a more recent but smaller (surgery group: n = 25; non-surgery group: n = 59) study by Walsh et al. did not show statistical differences in total tissue volume or global brain abnormality scores between surgery and non-surgery group^14^. Most of the surgeries in their study were for PDA ligation (60%) and retinopathy of prematurity (32%). The differences in the findings between studies may be due to different sample sizes and etiologies requiring surgery. Our finding was also consistent with term infants who had neonatal noncardiac surgery. A case-control study showed surgery group had worse global brain abnormality scores and smaller biparietal diameters than the controls^28^. One study compared brain MRI findings after neonatal surgery for noncardiac congenital anomalies between term and preterm infants. Preterm infants who had neonatal noncardiac surgery more frequently had parenchymal lesions than term infants with noncardiac surgery^29^.

A bigger sample size than our study may be required to show an association between surgery exposure and worse neurodevelopmental outcomes. A large multi-center cohort study evaluating 12,111 very preterm infants (2,186 infants underwent surgery under general anesthesia) showed an association between exposure to surgery and neurodevelopmental impairment at two years of age after multivariable regression analysis^30^. In contrast, a study with a smaller sample size^13^ showed an association between surgery and neurodevelopmental impairment in a univariate analysis but lost statistical significance in multivariate analysis, like our current study. Another potential explanation for the lack of association between surgery exposure and abnormal neurodevelopment is that our participants received less extensive surgeries. Prior studies showed more surgery^31^ and longer surgery durations^14^ were associated with worse neurodevelopmental outcomes. However, this explanation is not supported because our participants likely received similar or potentially more extensive surgery than in prior studies because the total duration of surgery for our participants (median: 187 minutes) was longer than in prior studies (80 minutes^31^ and 148 minutes^14^).

Worse global brain abnormality scores in term brain MRI were associated with worse neurodevelopmental outcomes at two years, independent of exposure to surgery and clinical and social risk factors. This finding is consistent with recent studies in very preterm infants^32^ and extremely preterm infants^33^, although whether these studies included infants exposed to surgery under general anesthesia or not is unclear. Browers et al^33^ studied 239 extremely preterm infants and showed global brain abnormality score at term was associated with cognitive, fine motor, and gross motor scores at two years using multivariable regression analysis. A smaller cohort of very preterm infants also showed the association of global brain abnormality score at term with cognition, motor, and behavioral outcomes at two years^32^. Furthermore, the global brain abnormality score was associated with motor skills and behavior in the same cohort of very preterm infants at ten years old^32^. Our findings and other studies suggest the term brain MRI provides valuable information in predicting short-term and potentially longer-term neurodevelopmental outcomes of very preterm infants, including those who underwent surgeries under general anesthesia.

Our study has several limitations. First, 54 infants (14%), including five in the surgery group, did not have Bayley data. Speculating how this loss affected our results is challenging because these infants had favorable clinical profiles (higher gestational age, lower CRIB-II scores) but adverse social backgrounds (fewer private insurance). Second, as in many other prior studies, we did not have a pre-surgical brain MRI to ascertain the association between brain lesions and surgical exposure. Third, our sample size for different surgeries was not adequate to determine the effects of specific surgeries on brain MRI or neurodevelopment. Lastly, our neurodevelopmental outcomes were limited to 2 years for this study. We are following our study cohort to collect neurodevelopmental outcomes at school age. Despite these limitations, we believe our study results are robust because it is one of the largest imaging cohort studies of very preterm infants. Our study has an advantage in generalizability because it is a regional cohort study, including participants from academic and community NICUs in the region with few exclusions. Antenatal, perinatal, and postnatal data were acquired prospectively. Clinical and demographic data were collected using strict protocols and procedures, and MRI acquisition and processing algorithms were designed to mitigate ascertainment bias. To address confounding, we used propensity score analysis, a rigorous method to control for potential confounding.

## CONCLUSION

In this cohort, very preterm infants exposed to surgery under general anesthesia during their NICU stay were at risk of worse structural brain MRI measures at term-equivalent age independent of social and clinical risk factors. Exposure to surgery was not associated with worse neurodevelopmental outcomes at two years after controlling for clinical and social risk factors. The global brain abnormality score on MRI at term in very preterm infants was associated with neurodevelopmental outcomes at two years, independent of exposure to surgery or other confounding factors. Such brain MRI measures may serve as sensitive and early biomarkers of neurodevelopmental insults following major surgical interventions in very preterm infants and thus help identify the highest risk infants.

## Data Availability

All data produced in the present study are available upon reasonable request to the authors.

## ACKNOWLEDGEMENT

We sincerely thank the parents of infants that participated in our study and the Cincinnati Infant Neurodevelopment Early Prediction Study (CINEPS) Investigators: Principal Investigator: Nehal A. Parikh, DO, MS. Collaborators (in alphabetical order): Mekibib Altaye, PhD, Anita Arnsperger, RRT, Traci Beiersdorfer, RN BSN, Kaley Bridgewater, RT(MR) CNMT, Tanya Cahill, MD, Kim Cecil, PhD, Kent Dietrich, RT, Christen Distler, BSN RNC-NIC, Juanita Dudley, RN BSN, Brianne Georg, BS, Meredith Glover, BS, Cathy Grisby, RN BSN CCRC, Lacey Haas, RT(MR) CNMT, Karen Harpster, PhD, OT/RL, Lili He, PhD, Scott K. Holland, PhD, V.S. Priyanka Illapani, MS, Kristin Kirker, CRC, Julia E. Kline, PhD, Beth M. Kline-Fath, MD, Hailong Li, PhD, Matt Lanier, RT(MR) RT(R), Stephanie L. Merhar, MD MS, Greg Muthig, BS, Brenda B. Poindexter, MD MS, David Russell, JD, Kari Tepe, BSN RNC-NIC, Leanne Tamm, PhD, Julia Thompson, RN BSN, Jean A. Tkach, PhD, Hui Wang, PhD, Jinghua Wang, PhD, Brynne Williams, RT(MR) CNMT, Kelsey Wineland, RT(MR) CNMT, Sandra Wuertz, RN BSN CCRP, Donna Wuest, AS, Weihong Yuan, PhD. We also greatly appreciate the support of our NICU fellows, nurses, and staff, and most importantly, all the study families that made this research possible.

